# Evaluation of mhGAP-HIG training outcomes across multiple domains among primary care physicians in three humanitarian contexts in Pakistan: A mixed-methods study

**DOI:** 10.64898/2026.01.22.26344671

**Authors:** Asma Humayun, Asma Nisa, Israr ul Haq, Arooj Najmussaqib, Noor ul Ain Muneeb, Syed Tahir Hussain Shah, Hazrat Ali Khan, Shahnawaz Zehri, Kamal Khan Mandokhail, Hidayatullah Kakar

## Abstract

**Background:** Humanitarian settings in Pakistan face major gaps in mental health service delivery, with limited specialist resources and a high burden of untreated conditions. The WHO mhGAP-Humanitarian Intervention Guidelines (mhGAP-HIG) aim to strengthen the capacity of primary care physicians (PCPs) to identify and manage common mental health conditions in humanitarian contexts. However, the evidence for multidimensional training outcomes remains limited. This study evaluated the effectiveness of mhGAP-HIG training to improve outcomes in terms of knowledge, skills, attitudes and confidence among PCPs across different humanitarian contexts in Pakistan.

**Methods:** As part of implementing an evidence-based digital MHPSS service model, seven mhGAP-HIG workshops were held between September and November 2025 across two provinces and a federal area facing humanitarian contexts. We conducted a quasi-experimental, mixed-methods evaluation where a total of 149 PCPs completed standardized assessments of knowledge (mhGAP-HIG), therapeutic skills (ENACT), attitudes (MICA-4), and confidence. Paired sample t-tests and ANCOVA were used to assess within-group and between-group changes, respectively. Quantitative feedback on training quality was obtained and qualitative reflections on learning experiences were evaluated using thematic analysis.

**Results:** A modest improvement in attitude and significant improvements across all other domains were observed. The participants from KP showed greater gains in knowledge, while participants from GB showed greater gains in skills, as compared to other provinces. Participants expressed a high level of satisfaction in all workshops, and reported improvements in knowledge, confidence, and ability to identify, manage and refer people with common mental health conditions.

**Conclusion:** Systematic efforts to adapt and implement mhGAP-HIG may lead to significant improvements across multidimensional competencies of PCPs, including their knowledge, skills, attitudes and confidence.

## INTRODUCTION

As is the case with many other countries in the Global South, Pakistan experiences a complex myriad of conflict, climate, and economic-induced challenges (Ali et al., 2024; Ngcamu, 2023; Saeed, 2023). While these stressors exacerbate countries’ mental health and economic burdens (Ni et al., 2024; Sharpe & Davison, 2021), evidence shows that investment in mental healthcare generates significant economic returns (World Bank Group, 2016). With scarce specialist resources and an underwhelming allocation of only 0.4% of the health budget for mental health, Pakistan grapples to meet the huge mental health burden (Alvi et al., 2023; Main Thompson & Saleem, 2025). Based on this evidence, the Ministry of Planning, Development & Special Initiatives (MoPD&SI) in Pakistan has prioritized mental health and psychosocial support (MHPSS) as part of its core development agenda (MoPD&SI, 2022).

As a strategic approach to strengthen existing services within constrained resources, the MoPD&SI developed a digital, multi-tiered, and scalable MHPSS service model (Humayun, 2021) Following the recommendation by the World Health Organization for low-resource settings, a specific tier has been dedicated to integrate mental healthcare at the primary care level in this model (World Health Organization, 2023a). For this purpose, training tools were developed to build the capacity of PCPs by contextualizing the mhGAP-Humanitarian Intervention Guide (Humayun et al., 2025), a manual widely used to build the capacity of non-specialists to identify and manage common mental health conditions in the humanitarian contexts of LMICs (Echeverri et al., 2018; International Medical Corps, 2022; Tarannum et al., 2019; World Health Organisation, 2021). These training tools were piloted in Khyber Pakhtunkhwa (KP) to train PCPs, with encouraging results of the pre-post evaluation of training in terms of knowledge gains (Humayun & Najmussaqib, 2025). These findings are consistent with other experiences of implementing mhGAP-HIG (Siriwardhana et al., 2016; Tarannum et al., 2019).

However, our study had some methodological and contextual limitations. As first-line providers in low-resource and humanitarian settings, PCPs are often required to make rapid decisions about responding to mental healthcare needs, frequently in the absence of specialist support. In such contexts, the effectiveness of mhGAP-HIG training cannot be understood solely in terms of knowledge acquisition, rather through its influence on a broader set of interrelated competencies that shape clinical behavior at the point of care. Others have also suggested that reliance on knowledge assessment overlooks the opportunity to analyze other competencies critical for effective delivery (Keynejad et al., 2018).

Beyond individual-level competencies, the broader implementation context also shapes the extent to which mhGAP-HIG training outcomes translate in practice (Queiroz et al., 2025). Differences in service availability, patient volume, supervisory structures, referral pathways, and sociocultural perceptions of mental illness can influence both baseline provider capacity and the extent to which training gains are realized. We also recognized gaps in the literature regarding the extent to which implementation of the mhGAP-HIG addresses contextual and systemic factors across diverse settings (Faregh et al., 2019; Gómez-Carrillo et al., 2020).

These considerations underscore two imperatives: to evaluate participants’ post-training competencies on more than one dimension, and to assess the effectiveness of these trainings in different contexts. In light of these evidence gaps, we designed a study to comprehensively evaluate the effectiveness of mhGAP-HIG training based on a range of measurable performance indicators spanning multiple competency domains; and across different humanitarian and healthcare contexts in Pakistan: provinces of KP and Balochistan; and the federally administrative area of Baltistan.

Although these entities face consistent humanitarian challenges, they differ significantly in geo-political, socio-cultural and developmental characteristics, as well as in the structure and functions of their healthcare systems and humanitarian needs. KP’s context is defined by recurring natural disasters, forced displacements, war on terror, and a history of hosting large Afghan refugee populations (Ishfaq et al., 2022; UNHCR, 2020; UNOCHA, 2022). In contrast, Balochistan is Pakistan’s largest and most underserved province facing long-standing underdevelopment of health infrastructure, extreme geographic isolation, and complex security and climate-related conditions (Imdad, 2023; Population Council, 2025). Meanwhile, the humanitarian needs in Baltistan are primarily environmental (climate change impacts) and logistical (constraints to healthcare access and service delivery) in nature (Asif, 2017; Department of Health Baltistan, 2019).

The healthcare context is also distinct, as KP has a relatively more developed healthcare system, with major tertiary hospitals in urban clusters and accessible primary healthcare facilities. Baltistan has a small but relatively functional health system; however, Balochistan faces a severe shortage and poorly accessible facilities, resulting in poor health outcomes (Asif, 2017; Humayun, 2024; Khan et al., 2025).

These considerations underscore the need to evaluate mhGAP-HIG training using multidimensional indicators (World Health Organization, 2017) that capture changes across key provider competencies, while also examining how training effects manifest across diverse humanitarian and healthcare contexts.

## METHODS

The objective of this study was to evaluate the effectiveness of the mhGAP-HIG (adapted for Pakistan) based on a range of measurable performance metrics: knowledge, skills, attitude and confidence across diverse humanitarian settings of Pakistan.

### Study design and setting

A quasi-experimental, pre-post evaluation design was used with a mixed methods approach. We used a full-cohort approach and included all participants who completed assessments in the analysis. We collected the quantitative data through structured online pre- and post-training assessments, while qualitative data comprised participants’ semi-structured feedback (structured and open-ended) on learning outcomes and training experience. The participants were drawn from three non-randomized regional groups representing diverse humanitarian contexts within Pakistan: KP, Balochistan, and Gilgit-Baltistan (GB).

In collaboration with respective provincial health departments and development partners, we conducted seven mhGAP-HIG training workshops, 2 each in Haripur and Kohat (KP), 2 in Quetta (Balochistan); and 1 in Skardu (GB) between September and November 2025. The target districts were selected by the respective departments of Health, based on the humanitarian context, availability of funds, and feasibility of conducting a pilot project for implementing mhGAP-HIG guidelines.

### The participants

We employed a purposive sampling strategy to register a total of 149 PCPs: KP (75), Balochistan (57), and Gilgit-Baltistan (17). The nominations of PCPs were provided by the respective health departments based on pre-defined inclusion criteria described previously (Humayun & Najmussaqib, 2025).

All PCPs registered on the MHPSS web-portal and completed both pre- and post-training assessments for knowledge, attitudes and confidence on the integrated Learning Management System (LMS), while skills were assessed through interactive role-play. All PCPs participated voluntarily after being informed of the study’s objectives and procedures. The consort figure for the study with the sample size is presented in *supplementary material A*. Informed verbal consent was obtained from all participants prior to data collection.

The demographic characteristics of participants are summarized in Table 1.

**Table 1.** Demographic details of PCPs (N = 149)

| **Table 1. Demographic details of PCPs (N = 149)** |  |  |  |
| --- | --- | --- | --- |
| **Demographic details** | **KP (N = 75)** | **Balochistan (N = 57)** | **GB (N = 17)** |
| Age (years), mean ± SD | 38.71 ± 6.62 | 36.86 ± 9.79 | 33.00 ± 7.91 |
| Gender, n (%) |  |  |  |
| Male | 44 (58.7%) | 39 (68.4%) | 11 (64.7%) |
| Female | 31 (41.3%) | 18 (31.6%) | 6 (35.3%) |
| **Years of experience, n (%)** |  |  |  |
| < 5 years | 15 (20.0%) | 22 (38.6%) | 15 (88.2%) |
| 5–10 years | 37 (49.3%) | 17 (29.8%) | 1 (5.9%) |
| > 10 years | 23 (30.7%) | 18 (31.6%) | 1 (5.9%) |
| **Educational qualification (PCPs), n (%)** |  |  |  |
| Medical degree (MBBS/FCPS/MD/MCPS) | 61 (81.3%) | 49 (86.0%) | 16 (94.1%) |
| Additional qualifications (FCPS/MD/MCPS) | 14 (18.7%) | 8 (14.0%) | 1 (5.9%) |
| **Health-care sector, n (%)** |  |  |  |
| Public sector | 72 (96.0%) | 50 (87.7%) | 12 (70.6%) |
| Private sector | 1 (1.3%) | 4 (7.0%) | 4 (23.5%) |
| Other / Not specified | 2 (2.7%) | 3 (5.3%) | 1 (5.9%) |

### The training workshops

The training workshops were led by a master trainer (AH) and a team of nationally certified mhGAP trainers (including IH, AN, STHS, HA, SZ, KKM, HK), focusing on the identification and management of common mental health conditions covered in mhGAP-HIG, adapted for humanitarian, cultural and healthcare context of Pakistan (Humayun et al., 2025). Each training workshop was conducted over five consecutive days, combining interactive and competency-based approaches such as large and small-group discussions, demonstrations, role-plays, hands-on skills practice, case studies and peer feedback, in accordance with the WHO mhGAP-HIG training methodology and facilitator guidelines (World Health Organization, 2023b). The training resources, structure, and delivery methodology of the mhGAP-HIG training programme in Pakistan have been described elsewhere (Humayun et al., 2025).

### The instruments

For a comprehensive evaluation of the effectiveness of the mhGAP-HIG training, we assessed four domains: knowledge, clinical competency (skills), attitudes and confidence. Knowledge, attitudes, and confidence were assessed using self-administered questionnaires, available in both English and Urdu languages, while skills were assessed through the ENACT tool. All assessments were conducted on the first and last day of the training. These are described as follows:

### The mhGAP-HIG Knowledge Assessment Questionnaire

We used the knowledge Questionnaire from the mhGAP-HIG facilitator’s guide (World Health Organization, 2022). It has two parts: 10 True/False statements and 15 multiple-choice questions. Higher scores indicated greater knowledge (range 0-26). The instrument has been translated and previously administered in mhGAP training evaluations in Pakistan and has demonstrated acceptable internal consistency (Humayun & Najmussaqib, 2025).

### Enhancing Assessment of Common Therapeutic Factors (ENACT)

To assess skills, we used an adapted version of the Enhancing Assessment of Common Therapeutic Factors (ENACT) tool, originally developed by Kohrt and colleagues (World Health Organization, 2021). The tool evaluates core therapeutic and communication skills, such as active listening, empathy, rapport building, and risk assessment.

Our main objective of adapting the ENACT tool for Pakistan was to produce a shorter version (based on eight ENACT competencies, instead of sixteen) as the original tool was deemed too resource-intense (Kohrt, Ramaiya, et al., 2015). The adaptation followed multiple expert consultations and focus group discussions to develop a scripted role play based on instructions for both participants and actors. Total scores ranged from 8 to 32, with higher scores indicating greater competency. The original ENACT scale demonstrated strong internal consistency in the initial validation study (Kohrt, Jordans, et al., 2015). The complete process of adaptation of ENACT for Pakistan is described elsewhere (Humayun et al., 2026).

### Mental Illness Clinicians’ Attitudes Scale - MICA-4

We used the Mental Illness Clinicians’ Attitudes Scale (MICA-4) to measure attitudes of PCPs towards people with mental health problems (Kassam et al., 2010). The MICA-4 comprises 16 items scored on a six-point Likert scale, with higher scores indicating more stigmatizing attitudes (range 16-96). In its original validation study, the MICA-4 demonstrated acceptable internal consistency (Gabbidon et al., 2013). For this study, the tool was translated and culturally adapted for use in the local context, and the validation process will be described elsewhere.

### The Confidence Questionnaire

We measured the confidence of PCPs using a confidence questionnaire developed for evaluating mhGAP training in non-specialized health settings (Wright et al., 2014), translated for use in our local context.

The tool includes 14 items rated on a four-point Likert scale, with higher scores reflecting greater confidence (range 14-56). The tool has been previously administered and tested in mhGAP training, with good internal consistency (Kokota et al., 2020).

### Feedback form

Feedback was collected using a digital form on LMS. The quantitative feedback covered key dimensions of training quality, including training resources, duration, logistics and planning, peer learning, overall experience, opportunity to participate, training methodology, and facilitator skills. Ratings were recorded on a 5-point Likert scale (1 = Unsatisfactory, 5 = Excellent). In addition, the feedback form included one open-ended question to capture participants’ qualitative comments on the training experience and suggestions for improvement.

### Data analysis

We entered, cleaned, and analyzed all data using IBM SPSS Statistics, version 29.0 (IBM, 2024).

Analyses were conducted using measure-specific analytical samples. Knowledge, attitudes, and confidence (K/A/C) outcomes were analyzed using all available data from participants who completed these assessments. For the ENACT competency measure, multiple imputation was used to address partially observed data (e.g., missing pre- or post-training scores), and to reduce bias associated with listwise deletion. Five imputations were generated, and pooled estimates are reported for all ENACT-related analyses.

Paired-sample *t* tests were used to examine pre–post changes in knowledge, clinical competency, attitudes, and confidence. Baseline differences between provinces were assessed using one-way analysis of variance (ANOVA), while between-province comparisons were assessed using analysis of covariance (ANCOVA). Bonferroni-adjusted post-hoc comparisons were applied where appropriate.

Effect sizes were estimated using Cohen’s *d* for pre–post comparisons and partial eta squared for covariance analyses. For qualitative feedback, descriptive statistics summarized the categorical feedback items, while responses to the open-ended question were analyzed thematically using Braun and Clarke’s six-step framework to explore participants’ perspectives on training experience (Braun & Clarke, 2006).

## RESULTS

Table 2 presents pre-post comparison of scores in terms of knowledge, attitude, confidence and skills.

**Table 2.** Knowledge, attitude, confidence, and ENACT comparisons pre- and post-training.

| Region | Domain | Pre M (SD) | Post M (SD) | *t* | *p* | Cohen's *d* |
| --- | --- | --- | --- | --- | --- | --- |
| **All trainings** | Knowledge | 16.88 (3.52) | 20.58 (3.13) | -12.03 | < .001 | -0.87 |
|  | Attitude | 44.88 (9.64) | 40.01 (10.70) | 5.67 | < .001 | 0.44 |
|  | Confidence | 38.66 (7.88) | 49.39 (5.39) | -15.85 | < .001 | -1.21 |
|  | ENACT | 1.69 (0.27) | 2.58 (0.32) | -31.53 | < .001 | -2.76 |
| **KP** | Knowledge | 17.70 (2.88) | 21.81 (2.34) | -10.52 | < .001 | -1.18 |
|  | Attitude | 43.13 (9.38) | 37.95 (10.23) | 4.89 | < .001 | 0.55 |
|  | Confidence | 36.38 (7.82) | 49.55 (5.81) | -14.25 | < .001 | -1.63 |
|  | ENACT | 1.72 (0.28) | 2.53 (0.31) | -23.18 | < .001 | -2.73 |
| **Balochistan** | Knowledge | 15.44 (3.88) | 19.11 (3.52) | -6.71 | < .001 | -0.90 |
|  | Attitude | 47.71 (9.88) | 43.47 (10.86) | 2.60 | .012 | 0.35 |
|  | Confidence | 41.61 (7.04) | 48.88 (4.90) | -7.43 | < .001 | -0.98 |
|  | ENACT | 1.72 (0.26) | 2.53 (0.31) | -26.45 | < .001 | -4.13 |
| **GB** | Knowledge | 17.75 (3.80) | 19.44 (2.68) | -1.84 | .085 | -0.46 |
|  | Attitude | 43.81 (8.25) | 38.25 (9.89) | 2.12 | .051 | 0.53 |
|  | Confidence | 39.00 (7.84) | 50.44 (5.11) | -5.52 | < .001 | -1.38 |
|  | ENACT | 1.54 (0.23) | 2.92 (0.12) | -27.21 | < .001 | -6.60 |

Significant improvements were observed in knowledge, skills and confidence across all provinces, while changes in attitudes toward mental illness were modest.

Baseline differences across provinces were observed for knowledge, skills, attitudes and confidence scores. Accordingly, analyses of covariance were used to compare post-training outcomes across provinces, adjusting for baseline scores (Table 3), while the post-hoc analyses are presented in *supplementary material B*.

**Table 3.**
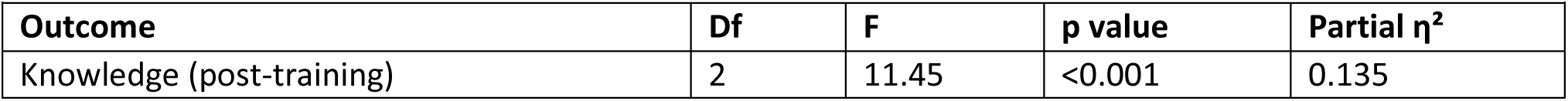
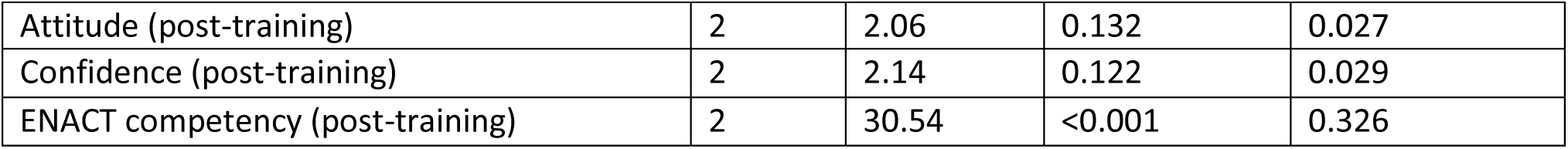
Between-province comparisons of post-training outcomes adjusted for baseline scores (ANCOVA)

| Outcome | Df | F | p value | Partial $\eta^2$ |
| --- | --- | --- | --- | --- |
| Knowledge (post-training) | 2 | 11.45 | <0.001 | 0.135 |
| Attitude (post-training) | 2 | 2.06 | 0.132 | 0.027 |
| Confidence (post-training) | 2 | 2.14 | 0.122 | 0.029 |
| ENACT competency (post-training) | 2 | 30.54 | <0.001 | 0.326 |

After adjustment for baseline scores, post-training knowledge and skills were significantly different across provinces. In contrast, no statistically significant provincial differences were observed for post-training attitudes or confidence. Pairwise comparisons of adjusted means showed significant differences in post-training ENACT scores, which were highest in GB (adjusted mean 3.02), compared with KP (2.51) and Balochistan (2.52). For knowledge, adjusted post-training scores were higher in KP (21.61) than in both Balochistan (19.47) and GB (19.22). These adjusted differences indicate greater post-training gains in skills among participants from GB and greater gains in knowledge among participants from KP, after accounting for baseline variation.

### Feedback

The quantitative feedback across all mhGAP training workshops showed that the participants rated high levels of satisfaction (either excellent or good) for the overall experience, training content, role of the facilitators, and teaching methodology. 10-15% participants shared reservations about the logistics and training duration rating these as fair or needs Improvement (see Figure 1).

**Figure 1:**
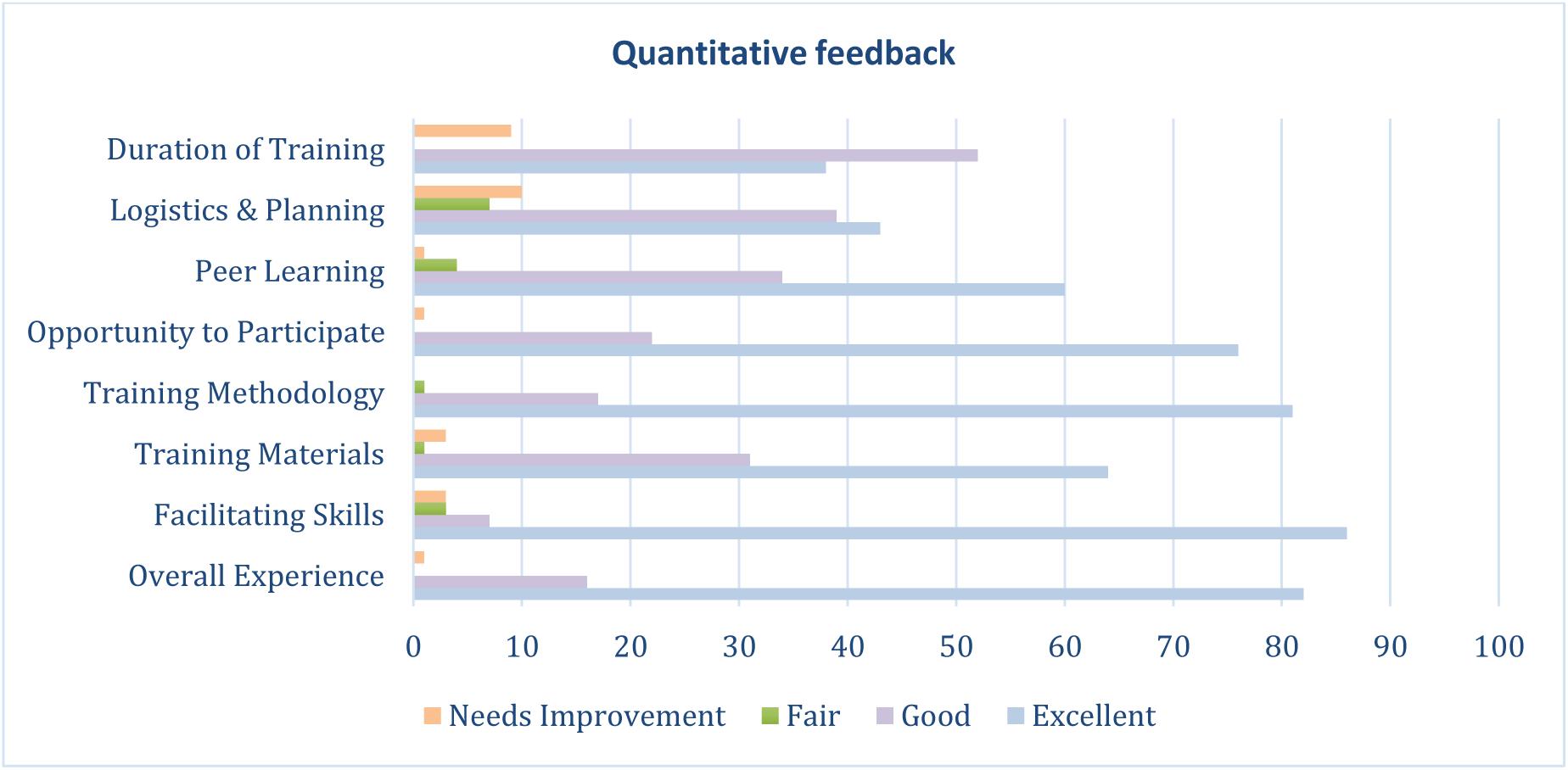
Quantitative feedback.

The qualitative feedback was also encouraging as the participants consistently emphasized that the mhGAP training strengthened their clinical capacity and confidence to identify and manage mental health conditions in primary-care practice. They valued the structured, interactive, and supportive learning environment, which helped foster collaboration across different levels of care. Suggestions for continuous supervision and refresher training emphasized the need for sustained efforts through ongoing institutional support to integrate mental healthcare within primary care.

Table 4 shows the thematic analysis of the participants’ feedback, illustrating perceived improvements in clinical competence and perspectives, and bridging the gap between theoretical understanding and applied confidence in routine service delivery, after the training.

**Table 4:** Thematic analysis of the participants’ feedback.

| Table 4: Thematic analysis of the participants' feedback |  |  |  |
| --- | --- | --- | --- |
|  | Themes/Subthemes | Description | Verbatim examples |
| **1** | **Clinical competence**<br>This theme captures participants' perceptions of how the mhGAP training enhanced their knowledge and skills through a systematic approach to help identify, manage, and follow up patients presenting with common mental health conditions within primary care settings. |  |  |
| 1.1 | To assess | Participants reported their practices of either dismissing people with mental disorders or instantly referring them to specialist services without attempting to assess the problem, as they believed that diagnosing mental disorders was an exclusive domain of psychiatrists. The training has equipped them with essential tools to take focused history, explore clinical presentations and recognize common mental health conditions to make assessment decisions. | *"Before, I was not able to diagnose patients with mental health conditions, but now I can assess properly."*<br>*"Earlier, I used to refer most cases thinking I couldn't manage them; now I can differentiate depression from normal sadness and initiate treatment."* |
| 1.2 | To manage | Participants recognized the training's role in bridging the gap between theoretical knowledge and clinical practice, particularly to help them manage people with mental health concerns. They described that case-based discussions and role play practice sessions as essential for translating knowledge into practice. These methods provided opportunities to rehearse communication techniques, practice psychosocial interventions, and make management decisions. | *"This workshop was very practical and helpful for managing patients."*<br>*"The training session was so much informative... learned so many things that I can apply in my clinical work."*<br>*"Very experienced, hands-on training... now I can deal better with psychiatric patients."* |
| 1.3 | To refer | Participants reported a clear understanding of which conditions can be managed in primary care and when specialist care becomes necessary. The training helped them determine the level of intervention required, the *Dos and Don'ts* of their responsibilities, and the need to work closely with specialist services. This indicates a shift in their mindset, from spontaneous practices to refer toward a proactive clinical involvement to assess and manage cases where possible. | *"Now I know when to treat and when to refer the patient ahead."*<br>*"The training has been very helpful and informative to manage patients on daily basis at primary care facilities. Now I understand when a follow-up is needed within primary care and when specialist support is indicated."*<br>*"I have more clarity on which patient needs treatment and which patient needs a referral."* |
| **2** | **Training programme**<br>This theme includes the feedback related to the planning, design and logistics of training workshops. |  |  |
| 2.1 | Role of trainers | Participants appreciated the crucial role of trainers in creating a respectful, inclusive, and engaging learning environment. | *"I liked the friendly atmosphere; it made learning easy."* |
|  |  | They described the trainers as approachable and responsive to learner needs; and highlighted their ability to simplify complex concepts and maintain participants' interest throughout the sessions. These factors enhanced participants' confidence to ask questions, encouraged open discussion to clarify uncertainties, and reduced the hesitation often associated with addressing mental-health topics in primary care. | *"The facilitators created a supportive and respectful learning environment where all participants felt comfortable to share and ask questions."*<br>*"They explained complex concepts clearly and used practical examples that made the material easy to understand."* |
| 2.2 | Teaching methodology | Participants highlighted the value of interactive and practice-oriented teaching methods that bridged theoretical learning with real-life clinical application. They regarded role-plays, case-based discussions, and group activities as essential elements that enhanced understanding, retention, and confidence; and enabled them to translate mhGAP principles into practical communication, assessment, and decision-making skills.<br>The participatory approach was particularly effective in maintaining focus and learn through reflection and peer experiences. | *"Role plays really helped to understand how to approach patients."*<br>*"Case discussions were very useful in remembering the important points."*<br>*"It was very practical training; we learned how to deal with patients in real situations."*<br>*"Interactive sessions kept us engaged throughout the training."* |
| 2.3 | Logistic and planning | The workshops were generally described as well-organized but highlighted some logistical gaps related to the venue arrangements and timing of the workshops. The latter was discussed by participants who had to travel long distances, sometimes over an hour, to attend the workshop. Some participants also found it challenging to balance attendance with clinical duties, suggesting shorter sessions to reduce fatigue, while others felt the existing duration was justified given the value of discussions. | *"Scheduling was difficult during duty hours."*<br>*"The duration should be minimized to three days."*<br>*"Five days were worth it; refresher courses should be held quarterly."*<br>*"The admin staff at the venue were a bit slow, but sessions and discussions were very good."*<br>*"Food quality and stationery need improvement."* |
| 3 | **Clinical perspective**<br>The training helped foster empathetic, inclusive, and patient-centered approaches while reducing hesitation and stigma. Participants reported feeling confident, motivated, and a shift in their mindset from a passive role to active engagement with people facing mental health challenges, thus developing a different outlook toward mental health care. |  |  |
| 3.1 | Collaborative paradigm | The training was described as a collaborative and engaging experience that emphasized teamwork and fostered a sense of partnership between primary care providers and specialists to improve patient outcomes. Many participants reflected that the experience reduced their hesitation to consult mental health professionals, making the process of managing mental health caseload more coordinated and less fragmented, thus bridging the disconnect that exists between primary and specialist care. | *"Good opportunity for GPs to learn and enhance their skills. Such trainings are really needed for patient welfare."*<br>*"It created understanding between doctors and specialists to work together."*<br>*"We learned from each other through participation and shared cases."* |
| 3.2 | Attitude and confidence | Participants described a marked shift in their perceptions and attitudes toward individuals experiencing mental health problems. Before the mhGAP training, several reported uncertainty, discomfort, or avoidance when dealing with such cases. After the training, many expressed a more empathetic and non-judgmental approach; a reduction in stigmatizing attitudes - both internal and systemic; and a shift from fear and misconceptions towards confidence and compassion. Participants shared a renewed sense of responsibility toward understanding patients' emotional and social contexts rather than labeling or dismissing them. | *"I personally found this training highly informative, and it enhanced my understanding... de-stigmatization of mental illness."*<br>*"Now I feel more comfortable talking to patients with mental illness."*<br>*"Before training I used to avoid such cases, now I can listen to them with patience."*<br>*"I feel more confident now in managing mental health patients that come to us first."* |
| 3.3 | Shift towards evidence-based practice | The mhGAP training prompted critical reflection on current practices and highlighted the need to uphold patient dignity and rights. Several participants acknowledged that individuals with mental illness are often marginalized, stigmatized, or maltreated. Following the training, participants emphasized the need to adopt evidence-based, ethically grounded approaches when working with individuals experiencing mental health problems. Through structured discussions and case-based examples, participants recognized that ethical care involves not only correct diagnosis and treatment but also respectful communication, confidentiality, and informed decision-making. | *"We have clearly understood the DON'TS: don't send patients to rehabilitation centers, don't prescribe benzodiazepines, and do not judge individuals with mental health problems".*<br>*"The training not only helped in improving our knowledge about mental health problems but also helped us to serve patients better. Such trainings are much needed for patient welfare."* |
| **4** | **Recommendations for integrating mental health in primary healthcare**<br>Participants highlighted the need for sustained efforts to strengthen mental health care integration within primary care settings through training more PCPs, continuous professional supervision, and structured refresher training. |  |  |
| 4.1 | Recognition of mental health care needs | Participants became more cognizant of the mental health burden in their communities and stressed that primary-care providers must actively respond to these unmet needs. They emphasized that mental health care is an essential component of holistic patient management and how they can help improve access to care.<br>These insights underscore the need for extended initiatives to build the capacity of primary healthcare providers and foster a multidisciplinary approach towards mental healthcare. | *“Really appreciate the efforts of the team... such trainings are really needed for patient welfare.”*<br>*“I suggest to conduct this type of training programs at district level also.”*<br>*“In my opinion, we should encourage awareness and encourage trainings to different healthcare workers apart from doctors as well so that it could be a multidisciplinary participation.”* |
| 4.2 | Pre-service training, ongoing supervision and continuous professional development | Participants repeatedly expressed their views that mhGAP training should not be viewed as a one-time event but as part of a continuous professional learning process. They strongly advocated structured refresher sessions, pre-service inclusion in medical curricula, and accessible supervision mechanisms to support sustained implementation of mhGAP guidelines. | *“It was a great learning opportunity... I wish to attend more sessions like this in future, and I hope you people arrange more sessions like this one.”*<br>*“This training should be included in undergraduate medical training.”* |

## DISCUSSION

Our mixed-methods evaluation study enriches the literature by offering insights into implementing mhGAP-HIG in three distinct humanitarian contexts across Pakistan. It offers a holistic evaluation of competencies and operational insights into implementing a large-scale competency-oriented mhGAP-HIG programme in resource-constrained public sector settings, facing humanitarian challenges.

Use of contextualized tools and multidimensional evaluation not only enhances methodological rigor but also offers in-depth reflections by the participants to explain improvements in their competencies. Embedding assessments within a scalable MHPSS digital model enhances its feasibility and scalability, while reducing administrative burden. Aligned with WHO guidelines, we did not rely on self-reported measures only, but also focused on observable competencies for a person-centered approach (Michael et al., 2020). We have also offered insights for quality improvement for future capacity-building initiatives to bridge the identified knowledge-to-practice gaps (Humayun et al., 2025).

The convergence between quantitative improvements and qualitative reflections indicates that PCPs not only assimilated knowledge but also developed perceived competence to apply skills in a person-centered manner. Consistent with recent scoping evidence from fragile and conflict-affected settings, qualitative feedback proved critical for identifying gaps in functional skills, risk assessment, and psychosocial engagement that are not captured by knowledge tests alone (Queiroz et al., 2025).

As proposed (Böhm et al., 2022), we moved beyond a didactic, manual-focused pedagogy to a competency-focused experiential learning through role-plays, simulations, structured feedback, and supervised case discussions. Evidence from Nigeria, Malawi, and Turkey also showed that didactic teaching alone is insufficient to change clinical practice (Adewuya et al., 2025; Karaoğlan Kahiloğulları et al., 2020; Kokota et al., 2020).

Hitherto, the mhGAP trainings have been evaluated within a single domain, most often knowledge, or by combining two domains (Aldana López et al., 2024; Asher et al., 2021; Karaoğlan Kahiloğulları et al., 2020; Kokota et al., 2020). It has also been suggested that improvement in knowledge might not be the best predictor for change in practices, as a positive association has been shown between improvement in attitudes and confidence and desirable practice patterns of patient engagement (Adewuya et al., 2025; Queiroz et al., 2025; Spagnolo & Lal, 2021).

Our findings demonstrated improvement in all domains, for instance, we observed an improvement in empathy and person-centered perspective in PCPs, but showed a reduction in negative attitudes and confidence to manage mental health problems within PHC, rather than referring them to the specialists. Based on our experience, we strongly agree that knowledge acquisition alone is insufficient for safe and effective task-sharing, and that overall competence and confidence of the PCPs must also be addressed so that they do not act primarily as gatekeepers for referral but actually function as first-line providers of mental health care. Others have also found that trainees improved on knowledge tests, yet struggled with psychoeducation, mental status examination, and risk assessment (Tarannum et al., 2019), or were not confident to deliver psychosocial interventions despite knowledge gains (Petagna et al., 2023).

We agree with Kokota and colleagues (2020) that despite a change in knowledge and confidence among primary care workers, a change in their attitude is much more difficult. As observed in Malawi (Ahrens et al., 2020), despite improvement in knowledge and confidence, there was no significant difference in terms of attitude in Balochistan and GB. This might be attributed to long-standing beliefs and prevailing stigma in primary care towards mental health (Husain et al., 2020). Also, 5-day training may not be enough to change attitude, and continuous supervision is needed so that physicians may not forget the training content or revert to their default attitude (Hana et al., 2024). Our study also observed province-wise change in competence of physicians, which might be attributable to variable pre-service education, mandating structured training programs at undergraduate and post-graduate levels (Siddiqui, 2018).

There is emerging evidence to use ENACT or ENACT-derived tools to assess communication and therapeutic skills as a component of mhGAP training (Kohrt et al., 2018, 2025; World Health Organization, 2025b).The challenges of implementing ENACT in resource-limited settings have also been reported (Kohrt, Jordans, et al., 2015; Mwenge et al., 2022; Spedding et al., 2022). We addressed these challenges by adapting it into a structured and brief measure encompassing eight core competencies to overcome its stated challenges in terms of resources, feasibility, cultural sensitivity, and administration (Humayun et al., 2026).

We understand that small changes in competencies may not be best assessed immediately after training (Jordans et al., 2022). We agree with Asher et al. (2021), that repeated ENACT assessments can be used to identify gaps in training programmes and that it remains to be seen whether the gains in these competencies would be sustained over time, and may best be assessed post-refresher training or during supervision.

Despite structured debriefing on the objectives of the assessments, it was observed by trainers that some PCPs were concerned that their evaluations might have professional implications. This underscores the importance of creating psychologically safe training spaces where participation is understood as formative and developmental, rather than purely assessment-driven. Similar dynamics have been reported by Tarannum et al., (2019) in humanitarian settings, where capacity building efforts may appear to challenge professional autonomy or established hierarchies, particularly in contexts influenced by organizational, gender, or seniority differences.

The need to carefully recruit PCPs eligible for mhGAP programmes has been previously highlighted (Humayun & Najmussaqib, 2025). Despite providing a pre-defined criterion, many participants were nominated by the district administration based on convenience. As a result, marked difference in the interest, engagement, and commitment towards delivering mental healthcare was observed. Similar observations were reported in Rohingya, Sub-Saharan African, and Nigeria’s mhGAP programmes, reinforcing the need for criteria-based recruitment (Adewuya et al., 2025; Jervase et al., 2022; Tarannum et al., 2019).

Based on our experience, career incentives should also be provided by the relevant health departments to encourage interested PCPs to provide mental healthcare.

We agree with Keynejad (2021) that the selection, motivation, and training of trainers is critical for engaging the PCPs and creating a conducive learning environment, and believe that effective simulation-based pedagogy requires facilitators who can model empathic communication, guide structured feedback, manage group dynamics, and apply competency scoring reliably.

Consistent with findings from mhGAP implementation in other humanitarian settings (Böhm et al., 2022; Tarannum et al., 2019), we also found that systemic constraints such as work pressures, high patient volumes, travel inconveniences, and overlapping administrative responsibilities influenced participation and engagement. Yet the overall consistency of improvements across provinces in our study demonstrates that meaningful gains are achievable despite logistic constraints.

We are cognizant that our study has yet not established sustained gains in knowledge, skills attitude, confidence; and plan to incorporate longitudinal monitoring into our programmes.

### Future implications

Current evidence strongly supports scaling up of mhGAP-HIG implementation to strengthen primary healthcare services across regions facing humanitarian contexts. Training programmes must prioritize competency-based learning outcomes and attitudinal transformation, with emphasis on communication and therapeutic skills, and efforts to reduce stigma. Global evidence increasingly supports shifting mhGAP implementation benchmarks from training completion toward demonstration of minimum competency thresholds, supported by ongoing supervision and refresher cycles (Queiroz et al., 2025).

Structured supervision, even if remote, and refresher cycles of training should be institutionalized as a routine element of service delivery (Gureje et al., 2015). This will also help develop effective working connections between the specialists and primary healthcare providers. These findings also emphasize the need to strengthen services at the primary care level and address systemic challenges related to work environments, managerial support, medication supply, and referral mechanisms. Pre-service integration of mhGAP competencies into medical, nursing, and psychology curricula is critical for effective integration of mental healthcare as a core function of the primary healthcare (World Health Organization, 2025a). Lastly, it is also essential to redefine the role of specialists and institutionalize their training as trainers and supervisors of PCPs for implementing mhGAP guidelines.

## Conclusion

This evaluation provides strong evidence that contextualized, competency-focused mhGAP-HIG training can significantly strengthen PCPs’ ability to identify and manage mental health conditions in Pakistan. By addressing critical gaps identified in global mhGAP literature, particularly in knowledge, attitudes, skills and confidence, this model offers a scalable and context-sensitive framework suitable for national scale-up in low-resource humanitarian settings aiming to integrate mental health into primary care in other LMICs as well.

## Data Availability

The data that support the findings of this study are available from the Health Section at the Ministry of Planning, Development & Special Initiatives, Government of Pakistan. Confidentiality restrictions apply to the availability of the data used under the license for the current study, and so are not publicly available. However, the data can be made available from the authors upon reasonable request after formal approval from the Ministry of Planning, Development & Special Initiatives, Pakistan.

## Authors Contribution

AH: Conceptualization, Methodology, Visualization, Supervision, Project administration, Writing – review and editing.

AsN: Methodology, formal analysis, data curation, Writing – original draft,

IH: Project implementation; Writing – original draft, data collection

AN: Methodology, data collection, project implementation

NM: Data collection, writing – original draft, project implementation

HA/THS/SZ/KHM/HK: Project implementation

## Acknowledgments

We would like to thank Mr Mujeeb ur Rehman, Secretary of Health Balochistan; Mr Asif ullah Khan, Secretary of Health, Gilgit-Baltistan; Dr Shahid Yunis, Director General Health Services, Khyber Pakhtunkhwa for extending their support.

We also acknowledge Deutsche Gesellschaft für Internationale Zusammenarbeit (GIZ) and International Medical Corps Pakistan; International Organization for Migration; and United Nation Population Fund (UNFPA), for their support to implement mhGAP-HIG in KP, Balochistan and Baltistan respectively.

## Ethical Statement

This study was conducted as part of the Mental Health and Psychosocial Support Project, approved by the Ministry of Planning, Development & Special Initiatives in compliance with ethical standards and consent protocols under letter no. 6(262) HPC/2020 and Health Department, KP under letter no. 2243-49/PH. Ethical approval was also sought from institutional review board of Bolan University under Approval No. 1078/IRB/BUMHS/25.

## List of abbreviations

PCPs: Primary care physicians
LMCs: Low-and-middle-income countries
mhGAP-HIG: mhGAP-Humanitarian Intervention Guidelines
ENACT: Enhancing Assessment of Common Therapeutic Factors
MICA: Mental Illness: Clinicians’ Attitudes Scale
MHPSS: Mental health and psychosocial support
KP: Khyber Pakhtunkhwa
GB: Gilgit Baltistan

## Supplementary material A

**Figure S1:**
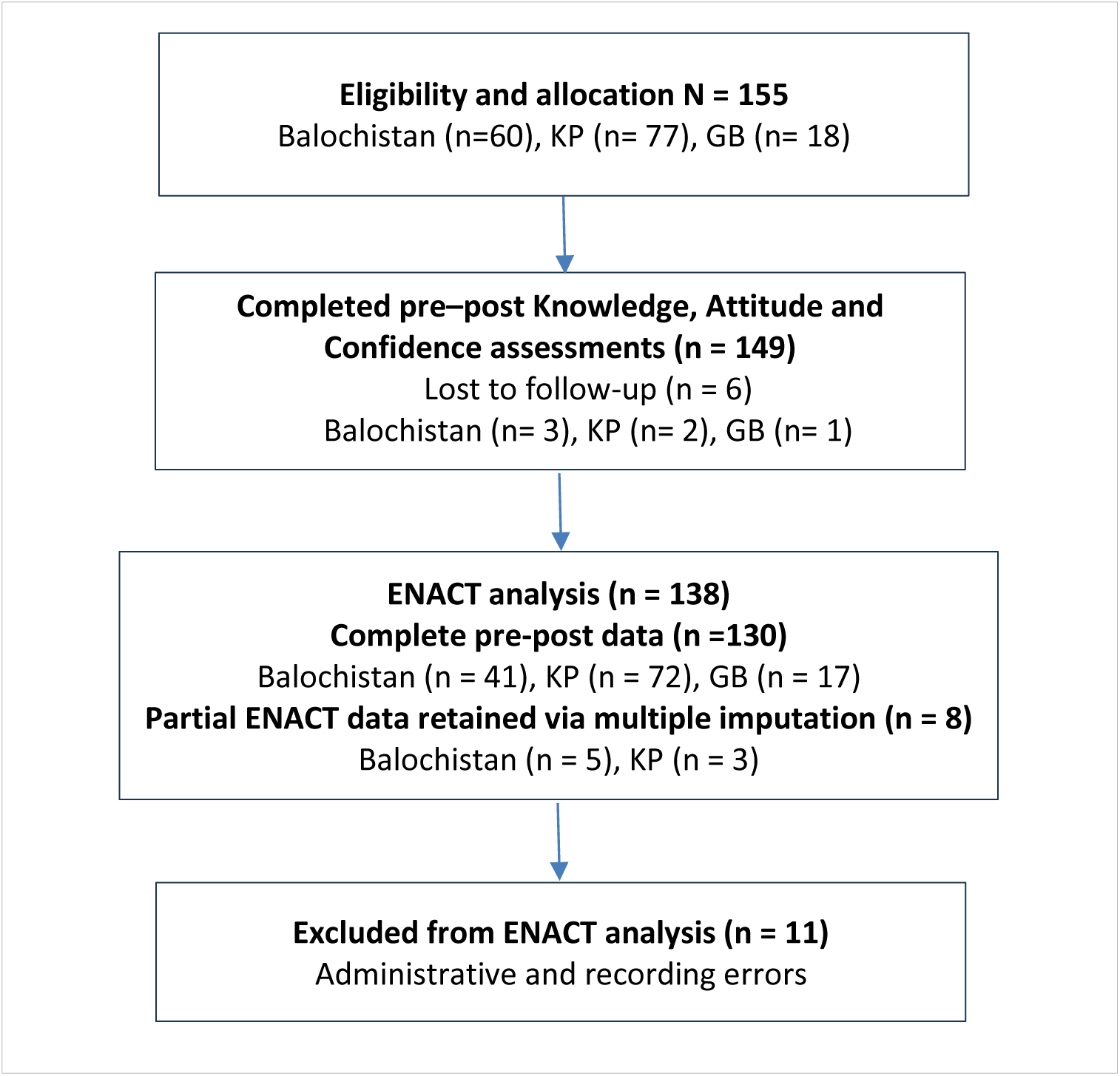
CONSORT flow diagram of the quasi-experimental study.

## Supplementary material B

**Table S1.** Post-hoc pairwise comparisons of baseline knowledge scores between provinces.

**Supplementary material B:**
|  |  |  |  |  |
| --- | --- | --- | --- | --- |
| **Province comparison** | **Mean difference** | **SE** | **95% CI** | ***p* (adjusted)** |
| Balochistan vs KP | 0.42 | 0.15 | [0.12, 0.72] | .006 |
| Balochistan vs Skardu | 0.18 | 0.17 | [-0.16, 0.52] | .294 |
| KP vs Skardu | -0.24 | 0.14 | [-0.52, 0.04] | .092 |

**Table S2.** Post-hoc pairwise comparisons of baseline attitude scores between provinces.

|  |  |  |  |  |
| --- | --- | --- | --- | --- |
| **Province comparison** | **Mean difference** | **SE** | **95% CI** | ***p* (adjusted)** |
| Balochistan vs KP | 0.31 | 0.13 | [0.05, 0.57] | .019 |
| Balochistan vs Skardu | 0.09 | 0.15 | [-0.21, 0.39] | .548 |
| KP vs Skardu | -0.22 | 0.12 | [-0.46, 0.02] | .067 |

**Table S3.** Post-hoc pairwise comparisons of baseline confidence scores between provinces.

|  |  |  |  |  |
| --- | --- | --- | --- | --- |
| **Province comparison** | **Mean difference** | **SE** | **95% CI** | ***p* (adjusted)** |
| Balochistan vs KP | 0.47 | 0.14 | [0.19, 0.75] | .002 |
| Balochistan vs Skardu | 0.21 | 0.16 | [-0.11, 0.53] | .184 |
| KP vs Skardu | -0.26 | 0.13 | [-0.52, -0.01] | .041 |

**Table S4.** Post-hoc pairwise comparisons of baseline ENACT total scores between provinces.

|  |  |  |  |  |
| --- | --- | --- | --- | --- |
| **Province comparison** | **Mean difference** | **SE** | **95% CI** | ***p* (adjusted)** |
| Balochistan vs KP | 0.34 | 0.11 | [0.12, 0.56] | .003 |
| Balochistan vs Skardu | 0.15 | 0.13 | [-0.11, 0.41] | .251 |
| KP vs Skardu | -0.19 | 0.10 | [-0.39, 0.01] | .061 |

## References

Adewuya, A., Ola, B., Coker, O., Atilola, O., Olibamoyo, O., Oladipo, O., & Ajomale, T. (2025). Scaling mental health care in Nigeria: Impact of WHO mhGAP training under the MeHPriC program on knowledge, attitudes, and practices of primary health care workers in Lagos State – A pre-post mixed-methods study. Cambridge Prisms: Global Mental Health, 12, e83. 10.1017/gmh.2025.10040

Ahrens, J., Kokota, D., Mafuta, C., Konyani, M., Chasweka, D., Mwale, O., Stewart, R. C., Osborn, M., Chikasema, B., Mcheka, M., Blackwood, D., & Gilfillan, S. (2020). Implementing an mhGAP-based training and supervision package to improve healthcare workers’ competencies and access to mental health care in Malawi. International Journal of Mental Health Systems, 14(1), 11. 10.1186/s13033-020-00345-y

Aldana López, J. A., Serrano Sánchez, M. del R., Páez Venegas, N., Chávez Sánchez, A. V., Flores Bizarro, A. D., Blanco Sierra, J. A., Jarero González, C. A., & Carmona Huerta, J. (2024). Impact of a social media-delivered distance learning program on mhGAP training among primary care providers in Jalisco, Mexico. BMC Medical Education, 24(1), 965. 10.1186/s12909-024-05950-w

Ali, N., Butzbach, O. K., Katohar, H. A., & Afridi, H. I. (2024). Structural and External Barriers to Pakistan’s Economic Growth: Pathways to Sustainable Development. World, 5(4), 1120–1129. 10.3390/world5040056

Alvi, M. H., Ashraf, T., Naz, F., Sardar, A., Ullah, A., Patel, A., Kiran, T., Gumber, A., & Husain, N. (2023). Burden of mental disorders by gender in Pakistan: analysis of Global Burden of Disease Study data for 1990–2019. BJPsych Bulletin, 1–8. 10.1192/bjb.2023.76

Asher, L., Birhane, R., Teferra, S., Milkias, B., Worku, B., Habtamu, A., Kohrt, B. A., & Hanlon, C. (2021). “Like a doctor, like a brother”: Achieving competence amongst lay health workers delivering community-based rehabilitation for people with schizophrenia in Ethiopia. PLOS ONE, 16(2), e0246158. 10.1371/journal.pone.0246158

Asif, A. F. (2017). Healthcare Challenges in Gilgit Baltistan: The way Forward. Pakistan Journal of Public Health, 7(2). https://pjph.org/pjph/article/view/47/35

Böhm, B., Palma, M., Ousley, J., & Keane, G. (2022). Competency-based mental health supervision: evidence-based tool needs for the humanitarian context. Global Mental Health, 9, 221–222. 10.1017/gmh.2022.23

Braun, V., & Clarke, V. (2006). Using thematic analysis in psychology. Qualitative Research in Psychology, 3(2), 77–101. 10.1191/1478088706qp063oa

Department of Health Baltistan. (2019). Health Sector Strategy Gilgit Baltistan (2013-2018). https://phkh.nhsrc.pk/sites/default/files/2019-06/HealthSectorStrategyGB2013-18.pdf

Echeverri, C., Le Roy, J., Worku, B., & Ventevogel, P. (2018). Mental health capacity building in refugee primary health care settings in Sub-Saharan Africa: impact, challenges and gaps. Global Mental Health (Cambridge, England), 5, e28. 10.1017/gmh.2018.19

Faregh, N., Lencucha, R., Ventevogel, P., Dubale, B. W., & Kirmayer, L. J. (2019). Considering culture, context and community in mhGAP implementation and training: Challenges and recommendations from the field. In International Journal of Mental Health Systems (Vol. 13, Issue 1). BioMed Central Ltd. 10.1186/s13033-019-0312-9

Gabbidon, J., Clement, S., van Nieuwenhuizen, A., Kassam, A., Brohan, E., Norman, I., & Thornicroft, G. (2013). Mental Illness: Clinicians’ Attitudes (MICA) Scale—Psychometric properties of a version for healthcare students and professionals. Psychiatry Research, 206(1), 81–87. 10.1016/j.psychres.2012.09.028

Gómez-Carrillo, A., Lencucha, R., Faregh, N., Veissière, S., & Kirmayer, L. J. (2020). Engaging culture and context in mhGAP implementation: fostering reflexive deliberation in practice. BMJ Global Health, 5(9), e002689. 10.1136/bmjgh-2020-002689

Gureje, O., Abdulmalik, J., Kola, L., Musa, E., Yasamy, M. T., & Adebayo, K. (2015). Integrating mental health into primary care in Nigeria: report of a demonstration project using the mental health gap action programme intervention guide. BMC Health Services Research, 15(1), 242. 10.1186/s12913-015-0911-3

Hana, R. A., Heim, E., Cuijpers, P., Sijbrandij, M., Chammay, R. El, & Kohrt, B. A. (2024). Addressing “what matters most” to reduce mental health stigma in primary healthcare settings: a qualitative study in Lebanon. BMC Primary Care, 25(1), 427. 10.1186/s12875-024-02680-2

Humayun, A. (2021). *A rights-based digital solution for public mental healthcare in Pakistan*. Ministry of Planning, Development and Special Initiatives.

Humayun, A. (2024, January 26). MHPSS roadmap in KP. *DAWN Newspaper*.

Humayun, A., Muneeb, N. ul A., Najmussaqib, A., Haq, I. ul, & Asif, M. (2025). Bridging the gaps: Contextualizing the mhGAP Humanitarian Intervention Guide to implement in Pakistan. Preprint. 10.1101/2025.05.21.25327990

Humayun, A., & Najmussaqib, A. (2025). Implementing the mhGAP-HIG: the process and evaluation of training primary healthcare workers in Khyber Pakhtunkhwa, Pakistan. BJPsych International, 1–11. 10.1192/bji.2025.10059

Humayun, A., Nisa, A., ul Haq, I., Najmussaqib, A., & Muneeb, N. ul A. (2026). Adaptation and psychometric evaluation of the ENhancing Assessment of Common Therapeutic factors (ENACT) tool to build the capacity of primary care physicians in low-resource settings. Preprint. 10.64898/2026.01.20.26344430

Husain, M. O., Zehra, S. S., Umer, M., Kiran, T., Husain, M., Soomro, M., Dunne, R., Sultan, S., Chaudhry, I. B., Naeem, F., Chaudhry, N., & Husain, N. (2020). Stigma toward mental and physical illness: attitudes of healthcare professionals, healthcare students and the general public in Pakistan. BJPsych Open, 6(5), e81. 10.1192/bjo.2020.66

IBM. (2024). IBM SPSS Statistics 29. https://www.ibm.com/support/pages/downloading-ibm-spss-statistics-29

Imdad, I. (2023). The post-flood mental health crisis in Balochistan. *DAWN Newspaper*. https://www.dawn.com/news/1732645#:∼:text=Regrettably%2C the governmental and non,deterioration of their mental wellbeing.

International Medical Corps. (2022). Field Implementation of the 2021 mhGAP-HIG Capacity Building Project: A Case Study. https://cdn1.internationalmedicalcorps.org/wp-content/uploads/2022/06/Field-Implementation-of-the-2021-mhGAP-HIG-Capacity-Building-Project_A-Case-Study.pdf

Ishfaq, U., Ashfaq, K., & Haroon, M. (2022). War on Terror and its Implications on Khyber Pakhtunkhwa. Global International Relations Review, V(IV), 13–20. 10.31703/girr.2022(V-IV).02

Jervase, R., Adams, B., Myaba, J., & Vallières, F. (2022). An evaluation of mental health capacity building among Community Rehabilitation Officers in Malawi: A mixed-methods case study. SSM - Mental Health, 2. 10.1016/j.ssmmh.2022.100108

Jordans, M. J. D., Steen, F., Koppenol-Gonzalez, G. V., El Masri, R., Coetzee, A. R., Chamate, S., Ghatasheh, M., Pedersen, G. A., Itani, M., El Chammay, R., Schafer, A., & Kohrt, B. A. (2022). Evaluation of competency-driven training for facilitators delivering a psychological intervention for children in Lebanon: a proof-of-concept study. Epidemiology and Psychiatric Sciences, 31, e48. 10.1017/S2045796022000348

Karaoğlan Kahiloğulları, A., Alataş, E., Ertuğrul, F., & Malaj, A. (2020). Responding to mental health needs of Syrian refugees in Turkey: mhGAP training impact assessment. International Journal of Mental Health Systems, 14(1), 84. 10.1186/s13033-020-00416-0

Kassam, A., Glozier, N., Leese, M., Henderson, C., & Thornicroft, G. (2010). Development and responsiveness of a scale to measure clinicians’ attitudes to people with mental illness (medical student version). Acta Psychiatrica Scandinavica, 122(2), 153–161. 10.1111/j.1600-0447.2010.01562.x

Keynejad, R. C., Dua, T., Barbui, C., & Thornicroft, G. (2018). WHO Mental Health Gap Action Programme (mhGAP) Intervention Guide: A systematic review of evidence from low and middleincome countries. In Evidence-Based Mental Health (Vol. 21, Issue 1, pp. 29–33). BMJ Publishing Group. 10.1136/eb-2017-102750

Khan, H. A., Ashraf, S., Hasni, L., Anwar, M., Khalid, S. A., Sulaiman, M., Ullah, A., & Kakar, S. U. (2025). Access to Mental Health Services for Children in Balochistan. Biological and Clinical Sciences Research Journal, 6(6), 301–305. 10.54112/bcsrj.v6i6.1916

Kohrt, B. A., Asher, L., Bhardwaj, A., Fazel, M., Jordans, M. J. D., Mutamba, B. B., Nadkarni, A., Pedersen, G. A., Singla, D. R., & Patel, V. (2018). The Role of Communities in Mental Health Care in Low- and Middle-Income Countries: A Meta-Review of Components and Competencies. International Journal of Environmental Research and Public Health, 15(6), 1279. 10.3390/ijerph15061279

Kohrt, B. A., Jordans, M. J. D., Rai, S., Shrestha, P., Luitel, N. P., Ramaiya, M. K., Singla, D. R., & Patel, V. (2015). Therapist competence in global mental health: Development of the ENhancing Assessment of Common Therapeutic factors (ENACT) rating scale. Behaviour Research and Therapy, 69, 11–21. 10.1016/j.brat.2015.03.009

Kohrt, B. A., Pedersen, G. A., Schafer, A., Carswell, K., Rupp, F., Jordans, M. J. D., West, E., Akellot, J., Collins, P. Y., Contreras, C., Galea, J. T., Gebrekristos, F., Mathai, M., Metz, K., Morina, N., Mwenge, M. M., Steen, F., Willhoite, A., van Ommeren, M., … Yurtaev, A. (2025). Competency-based training and supervision: development of the WHO-UNICEF Ensuring Quality in Psychosocial and Mental Health Care (EQUIP) initiative. The Lancet Psychiatry, 12(1), 67–80. 10.1016/S2215-0366(24)00183-4

Kohrt, B. A., Ramaiya, M. K., Rai, S., Bhardwaj, A., & Jordans, M. J. D. (2015). Development of a scoring system for non-specialist ratings of clinical competence in global mental health: a qualitative process evaluation of the Enhancing Assessment of Common Therapeutic Factors (ENACT) scale. Global Mental Health, 2, e23. 10.1017/gmh.2015.21

Kokota, D., Lund, C., Ahrens, J., Breuer, E., & Gilfillan, S. (2020). Evaluation of mhGAP training for primary healthcare workers in Mulanje, Malawi: a quasi-experimental and time series study. International Journal of Mental Health Systems, 14(1), 3. 10.1186/s13033-020-0337-0

Main Thompson, A., & Saleem, S. M. (2025). Closing the mental health gap: transforming Pakistan’s mental health landscape. Frontiers in Health Services, 4. 10.3389/frhs.2024.1471528

Michael, S., Chowdhary, N., Rawstorne, P., & Dua, T. (2020). Developing competencies for the WHO mhGAP Intervention Guide Version 2.0 training package. World Psychiatry, 19(2), 248–249. 10.1002/wps.20762

MoPD&SI. (2022). 5 Es framework to turnaround Pakistan. https://www.pc.gov.pk/uploads/downloads/5Es_Framework.pdf

Mwenge, M. M., Figge, C. J., Metz, K., Kane, J. C., Kohrt, B. A., Pedersen, G. A., Sikazwe, I., Van Wyk, S. S., Mulemba, S. M., & Murray, L. K. (2022). Improving inter-rater reliability of the enhancing assessment of common therapeutic factors (ENACT) measure through training of raters. Journal of Public Health in Africa, 13(3), 12. 10.4081/jphia.2022.2201

Ngcamu, B. S. (2023). Climate change effects on vulnerable populations in the Global South: a systematic review. Natural Hazards, 118(2), 977–991. 10.1007/s11069-023-06070-2

Ni, C.-F., Lundblad, R., & Dykeman, C. (2024). Diversity and training delivery trends in psychological first aid during COVID-19: Implications for researchers and practitioners. *Psychological Trauma: Theory, Research*, Practice, and Policy, 16(2), 225–232. 10.1037/tra0001447

Petagna, M., Marley, C., Guerra, C., Calia, C., & Reid, C. (2023). Mental Health Gap Action Programme intervention Guide (mhGAP-IG) for Child and Adolescent Mental Health in Low- and Middle-Income Countries (LMIC): A Systematic Review. Community Mental Health Journal, 59(1), 192–204. 10.1007/s10597-022-00981-3

Population Council. (2025). District Vulnerability Index for Pakistan (DVIP): Harnessing multisectoral data to inform equitable policy and climate action. 10.31899/sbsr2025.1046

Queiroz, M. R. de, Rubini, E., Valente, M., Hubloue, I., & Corte, F. Della. (2025). Integrating mental health into primary health care in fragile and conflict-affected settings: a scoping review of mhGAP effectiveness. Cadernos de Saúde Pública, 41(9). 10.1590/0102-311xen199224

Saeed, R. (2023). Climate Change and Climate Induced Migration in Pakistan: A Threat to Human Security (The Study of RajanPur and Taunsa Sharif after Flood 2022). Annals of Human and Social Sciences, 4(IV). 10.35484/ahss.2023(4-IV)37

Sharpe, I., & Davison, C. M. (2021). Climate change, climate-related disasters and mental disorder in low- and middle-income countries: a scoping review. BMJ Open, 11(10), e051908. 10.1136/bmjopen-2021-051908

Siddiqui, S. Z. (2018). Medical education at crossroads: Recommendationsfrom a national study in Pakistan. Pak J Med Sci, 34(3), 772–775. https://www.researchgate.net/publication/26713366_Medical_Education_in_Pakistan

Siriwardhana, C., Adikari, A., Jayaweera, K., Abeyrathna, B., & Sumathipala, A. (2016). Integrating mental health into primary care for post-conflict populations: a pilot study. International Journal of Mental Health Systems, 10(1), 12. 10.1186/s13033-016-0046-x

Spagnolo, J., & Lal, S. (2021). Implementation and use of the Mental Health Gap Action Programme Intervention Guide (mhGAP-IG): A review of the grey literature. Journal of Global Health, 11, 04022. 10.7189/jogh.11.04022

Spedding, M., Kohrt, B., Myers, B., Stein, D. J., Petersen, I., Lund, C., & Sorsdahl, K. (2022). ENhancing Assessment of Common Therapeutic factors (ENACT) tool: adaptation and psychometric properties in South Africa. Global Mental Health, 9, 375–383. 10.1017/gmh.2022.40

Tarannum, S., Elshazly, M., Harlass, S., & Ventevogel, P. (2019). Integrating mental health into primary health care in Rohingya refugee settings in Bangladesh: experiences of UNHCR. Intervention, 17(2), 130. 10.4103/INTV.INTV_34_19

UNHCR. (2020). Pakistan Refugee Policy Review. http://www.sbp.org.pk/bprd/2019/CL2.htm.

UNOCHA. (2022). Detail Needs Assessment Flood Emergency-KP MEAL-Islamic Relief Pakistan. https://reliefweb.int/report/pakistan/detail-needs-assessment-flood-emergency-kp-25th-sep-2022

World Bank Group. (2016). Investing in treatment for depression and anxiety leads to fourfold return. https://www.worldbank.org/en/news/press-release/2016/04/13/investing-in-treatment-for-depression-anxiety-leads-to-fourfold-return

World Health Organisation. (2021). Story of change in four countries. In World Health Organization. https://www.who.int/publications/i/item/9789240037229

World Health Organization. (2017). mhGAP training manuals for the mhGAP Intervention Guide for mental, neurological and substance use disorders in non-specialized health settings, version 2.0 (for field testing). *Geneva*: https://iris.who.int/server/api/core/bitstreams/eaea9a17-08b5-43a1-984e-92babd0b7203/content

World Health Organization. (2021). ENhancing Assessment of Common Therapeutic factors (ENACT) competency assessment tool (English, in-person) (v1.0). https://equipcompetency.org/sites/default/files/downloads/2024-11/ENACT_inperson_published_240321.pdf

World Health Organization. (2022). mhGAP Humanitarian Intervention Guide (mhGAP-HIG) training of health-care providers: training manual. *Geneva*: https://www.unhcr.org/sites/default/files/legacy-pdf/6253f78c4.pdf

World Health Organization. (2023a). Integrating mental health in primary health care: part 1. The context for integration of mental health services in primary health care. Cairo: WHO Regional Office for the Eastern Mediterranean. https://applications.emro.who.int/docs/9789292740924-eng.pdf

World Health Organization. (2023b). Mental Health Gap Action Programme (mhGAP) guideline for mental, neurological and substance use disorders, 3rd edition. https://www.who.int/publications/i/item/9789240084278

World Health Organization. (2025a). Educating medical and nursing students to provide mental health, neurological and substance use care: A practical guide for pre-service education. https://www.who.int/publications/i/item/9789240104129

World Health Organization. (2025b). mhGAP Assessment & Management Behavioural Observation Competencies. Equip Competency, mhGAP v1.0. Equip Competency. https://equipcompetency.org/sites/default/files/downloads/2025-03/mhGAP_07032025.pdf

Wright, J., Common, S., Kauye, F., & Chiwandira, C. (2014). Integrating community mental health within primary care in southern Malawi: A pilot educational intervention to enhance the role of health surveillance assistants. International Journal of Social Psychiatry, 60(2), 155–161. 10.1177/0020764012471924

